# Diagnostic errors are common in acute pediatric respiratory disease: a prospective, single-blinded multicenter diagnostic accuracy study in Australian Emergency Departments

**DOI:** 10.1101/2021.07.06.21259221

**Authors:** Paul Porter, Joanna Brisbane, Jamie Tan, Natasha Bear, Jennifer Choveaux, Phillip Della, Udantha Abeyratne

## Abstract

**Background:** Diagnostic errors are a global health priority and a common cause of preventable harm. There is limited data available for the prevalence of misdiagnosis in pediatric acute-care settings. Respiratory illnesses, which are particularly challenging to diagnose, are the most frequent reason for presentation to pediatric emergency departments.

**Objective:** To determine the diagnostic error rate of acute childhood respiratory diseases in emergency departments.

**Methods:** Prospective, multicenter, single-blinded, diagnostic accuracy study in two well-resourced pediatric emergency departments in a large Australian city. Between September 2016 and August 2018, a convenience sample of children aged 29 days to 12 years who presented with respiratory symptoms was enrolled. The emergency department discharge diagnoses were reported by clinicians based upon standard clinical diagnostic definitions. These diagnoses were compared against consensus diagnoses given by an expert panel of pediatric specialists using standardized disease definitions after they reviewed all medical records.

**Results:** For 620 participants, the positive and negative percent agreement (%, [95% CI]) of the emergency department compared with the expert panel diagnoses were generally poor: isolated upper respiratory tract disease (61.4 [51.2, 70.9], 90.9 [88.1, 93.3]), croup (75.6 [64.9, 84.4], 97.9 [96.2, 98.9]), lower respiratory tract disease (86.4 [83.1, 89.6], 92.9 [87.7, 96.4]), bronchiolitis (66.9 [58.6, 74.5], 94.3 [80.8, 99.3]), asthma/reactive airway disease (91.0 [85.8, 94.8], 93.0 [90.1, 95.3]), clinical pneumonia (62·9 [49·7, 74·8], 95·0 [92·8, 96·7]), focal (consolidative) pneumonia (54·8 [38·7, 70·2], 86.2 [79.3, 91.5]). Only 59% of chest x-rays with consolidation were correctly identified. Between 6.9% and 14.5% of children were inappropriately prescribed based on their eventual diagnosis.

**Conclusion:** In well-resourced emergency departments, we have identified a previously unrecognized high diagnostic error rate for acute childhood respiratory disorders, particularly in pneumonia and bronchiolitis. These errors lead to the potential of avoidable harm and the administration of inappropriate treatment.

## INTRODUCTION

Diagnostic errors are the most common form of medical error and are defined as “the failure to make an accurate and timely explanation of the patient’s health problem or to communicate that explanation to the patient” (1). Although diagnostic errors are classified by the World Health Organization (WHO) as a global health priority and are regarded as a “moral, professional and public health imperative” by the U.S. National Academy of Medicine (1), there are a lack of published data and reliable measures in this area, particularly in child health.

Diagnostic error research has been hindered, in part, because clinicians are generally poor at recognizing their own mistakes, particularly if they have not been formally trained to identify errors (2-4). With these findings in mind, reported error rates are likely to be underestimates. In addition, physician confidence levels have been shown to be insensitive to their diagnostic accuracy or the case difficulty (5). In pediatrics, where a diagnosis is dependent on having an accessible and communicative caregiver, there could be even greater potential for error, which might contribute to the almost 7 million global childhood deaths each year (6). When surveyed, 15–77% of pediatricians reported making at least one diagnostic error a month, and 45% reported making at least one harmful error each year (7-9).

The consequences of diagnostic errors include preventable harm for individuals and cost to the public health sector. In the United States, an estimated one million people per year experience harm from misdiagnosis (10) with the potential level of harm being moderate to severe in up to 86% of cases (11,12). In total, the cost of medical errors is estimated to be USD 19.5billion per annum, with an economic impact approaching USD 1 trillion annually (13). Diagnostic errors are also an important contributor in medical malpractice (14).

Acute respiratory conditions are the most common reason for presentation to pediatric emergency departments (E.D.s) (15). However, there is little evidence in the literature to confirm the diagnostic accuracy of clinicians in these units. The differential diagnosis of childhood respiratory illness is challenging as it relies on a complex mix of clinical and interpretative skills in patient history, examination, and investigation. In the chaotic environment of a busy E.D., there is a greater potential for making errors. Many respiratory disorders have similar clinical features, such as breathlessness and wheeze, which can contribute to misdiagnoses (16). Auscultation underpins the diagnosis of many lower respiratory tract conditions, but this relies on clinician experience and interpretation and has been shown to have high inter-rater variation (17, 18). In primary care settings, respiratory diseases, including pneumonia and asthma, are commonly misdiagnosed; however, the error rates in E.D.s have not been reported (11, 19, 20). Due to the potential for serious harm arising from missing childhood respiratory diseases, there is a need to define how frequently errors occur in E.D.s. Once the breadth of the problem is defined, remedial approaches can be developed.

The present study aimed to determine the diagnostic error rates for acute childhood respiratory diseases in two well-resourced E.D.s in Western Australia.

## MATERIALS AND METHODS

### Ethics and study conduct

Approval to conduct the study was granted by the Human Research Ethics Committees of Ramsay Health Care WA|SA (Reference 1501) and the Child and Adolescent Health Service of Western Australia (Reference 2015030EP). We obtained written consent from the parents of all participants. No adverse events were reported. The study did not interfere with any aspect of clinical care or diagnosis.

### Study design and participants

This was a multicenter, prospective, single-blind diagnostic accuracy study. We recruited a convenience sample of children aged 29 days to 12 years who attended a study site with signs or symptoms of a respiratory illness (Box 1).

#### Box 1

Study inclusion and exclusion criteria

*Inclusion criteria* (at least one of the following)

- Rhinorrhea
- Cough
- Wheeze
- Stridor
- Increased work of breathing
- Shortness of breath

*Exclusion criteria*

- Lack of consent
- Mechanical ventilation (invasive, CPAP or BiPAP) or high-flow nasal cannula
- Too medically unstable to participate in the study as per treating clinician

Participating sites were two hospitals in Western Australia: a tertiary pediatric facility with 75,000 ED presentations per year and a metropolitan general hospital with 29,000 pediatric (109,000 total) E.D. presentations per year. Both hospitals are optimally resourced units with complete treatment, laboratory, and radiology services. The units deliver team-based clinical care led by Emergency Medicine Physicians and Pediatric and Emergency Fellowship registrars. At least two supervising consultants were always present in the departments.

## Data collection and definitions

### Collected data

We collected demographic data, clinical measures and symptoms, examination findings, cough-sound recordings, and diagnostic and treatment response reports. We recorded E.D. discharge diagnoses from hospital records, and diagnoses recorded by a consensus panel of pediatric specialists.

A research nurse recorded cough sounds onto a standard iPhone 6. Each participant provided five cough sound events (spontaneous or voluntary) whilst they were in the E.D.

### E.D. diagnosis

The E.D. diagnosis was recorded by the treating team at patient discharge from the unit, either to home or to an inpatient ward. The department’s clinicians reported x-rays performed in the E.D. The treating team could record more than one diagnosis for each participant. E.D. clinicians were blinded to the consensus panel diagnoses.

### Expert panel consensus diagnosis

To reach a consensus diagnosis, we assembled an expert panel comprising four acute-care pediatricians (Fellows of the Royal Australasian College of Physicians: median 15 years of specialist practice). All hospital charts and databases were available to the panel, including test results and treatment responses. The panel was able to access information related to the final discharge diagnosis from the E.D. and inpatient teams. Specialist radiologists reported all radiology, and the results were confirmed by the panel. For participants admitted to inpatient wards, data from their clinical course after the E.D. visit was available. The panel members were allowed to listen to the recorded coughs when considering a diagnosis of croup.

Two members of the panel reviewed each participant independently. A third member acted as a tiebreaker in the event of non-agreement. Each participant was scored as positive, negative, or unsure for each of the study conditions. A consensus diagnosis could not be assigned when there was insufficient information in the medical notes or if relevant tests were not performed.

## Study disease definitions

Study diagnostic definitions (Table 1) were developed from international guidelines and are consistent with the diagnostic pathways used at the two study sites (21-26) In addition to the specific diseases of “no respiratory disease found”, “croup”, “asthma/reactive airway disease” (asthma/RAD), “bronchiolitis” and “pneumonia”, we defined two broader groups to differentiate children with isolated upper respiratory disease (iURTD) from those with any form of lower respiratory tract disease (LRTD; disease below the trachea, including all chest infections and bronchodilator responsive conditions). Only children under 24 months were considered for a diagnosis of bronchiolitis (21, 25). The asthma/RAD group included children with acute asthma and viral-induced wheezy episodes that were bronchodilator responsive. A positive diagnosis of asthma/RAD required the participant to have a positive bronchodilator response documented by the clinical team during the treatment visit.

**Table 1:** Definition of study diagnoses.

| Disease | Essential features |
| --- | --- |
| Isolated upper respiratory tract disease (iURTD) | • Must meet both criteria:<br>○ Nasal congestion, rhinorrhea or a sore throat<br>○ No lower respiratory tract disease |
| Croup | • Must have: Typical ‘seal-like’ barking cough on the cough recording |
| Any lower respiratory tract disease (LRTD)<sup>a</sup> | • Any of the following:<br>○ Auscultatory findings (focal or generalized) including wheezing (or silent chest in the setting of severe obstruction), crackles, bronchial breath sounds, or decreased breath sounds<br>○ Increased work of breathing, unless purely associated with stridor (croup)<br>○ A productive cough >5 days (chronic bronchitis)<br>○ New consolidation, infiltrate or pleural effusion on chest x-ray |
| Bronchiolitis (Age <24 months only) | • Must have both:<br>○ A persistent cough<br>○ Diffuse wheeze that is non-responsive to bronchodilator<sup>b</sup> (if administered) or diffuse crepitations |
| Asthma/reactive airway disease (RAD) | • Must have both:<br>○ Wheeze (or silent chest in the setting of severe obstruction);<br>○ Documented responsiveness to bronchodilators<sup>b</sup> during this illness |
| Clinical Pneumonia | At least one feature from both of the following two categories:<br>○ History of either: (i) fever in prior 48 hours, (ii) cough, (iii) dyspnea, or (iv) chest pain<br>○ Auscultatory changes with either: (i) focal or generalized crepitations/wheeze, (ii) decreased breath sounds, or (iii) bronchial breathing |
| Focal (consolidative) Pneumonia | At least one feature from all the following three categories:<br>○ History of either: (i) fever in prior 48 hours, (ii) cough, (iii) dyspnea, or (iv) chest pain<br>○ A chest radiograph or computerized tomography scan showing either: (i) new consolidation or (ii) pleural effusion<sup>d</sup> (specialist radiologist report)<br>○ Auscultation revealed only focal (or no) abnormality. Bilateral, widespread wheeze or crepitations not allowed |

<sup>a</sup> LRTD is an umbrella term encompassing the diagnoses of any non-structural condition of the respiratory system with involvement below the level of the trachea. These include all chest infections and bronchodilator responsive conditions.
<sup>b</sup> Bronchodilator test — administration of salbutamol MDI (100µg per actuation) via spacer up to three times over 1 hour at the following doses: 6 inhalations for children ≤six years, 12 inhalations for children >6 years. No consensus diagnosis could be given for asthma/RAD or bronchiolitis (participant excluded) if a bronchodilator test was not administered (in participants >12 months of age.) The treating clinician must confirm all responses after the test is administered.
<sup>c</sup> Asthma/RAD includes conditions with bronchodilator responsiveness, including acute exacerbations of asthma and episodes of viral-induced, bronchodilator responsive wheeze.
<sup>d</sup> Radiology reported according to World Health Organisation standards to detect bacterial lung disease in post-vaccination surveillance programs (28,29).

Two sets of pneumonia were defined (27). The term “clinical pneumonia” was applied when radiology was not required in making the diagnosis, as is recommended in cases of mild severity by the Pediatric Infectious Diseases Society and the Infectious Diseases Society of America [27]. Focal (consolidative) pneumonia was examined as a separate group due to the likelihood of being bacterial in etiology. To be positive for focal (consolidative) pneumonia, a specialist radiologist report indicating new consolidation or pleural effusion was necessary along with only focal, or no, auscultatory findings. To standardize radiology interpretation for the expert panel, we used the WHO criteria for interpreting chest x-rays developed to report post-vaccination pneumonia prevalence (28, 29).

As was the case with the E.D. diagnosis, participants could have more than one study diagnosis as some were dependent subsets. For example, patients with pneumonia also received the broader diagnosis of LRTD.

### Statistical Analysis

As clinical diagnoses were considered non-reference standard measures, the primary measures of diagnostic agreement used were Positive Percent Agreement (PPA) and Negative Percent Agreement (NPA). 95% confidence intervals around these parameters were calculated using the method of Clopper-Pearson. When reporting demographic details, median and interquartile ranges are provided, given the skewed distribution.

All data were analyzed by an independent statistician using Stata 16.1 (StataCorp, College Station, Texas).

## RESULTS

### Demographics and expert panel diagnoses

We enrolled 620 children across the two study sites from September 2016 to August 2018: 125 at the tertiary pediatric hospital and 495 at the metropolitan general hospital. There were no differences in age (47 versus 47 months p=0.45) or sex (male: 60.2% versus 53.6%, p=0.18) between enrolment sites. The proportion of participants positive for each study diagnosis were similar at both sites, except for bronchiolitis, where more participants were recruited at the tertiary site (120/145 versus 25/39, p=0.016, data not shown). Participant flow through the study along with the reasons for any excluded cases per study disease is shown in figure 1.

**Figure 1:**
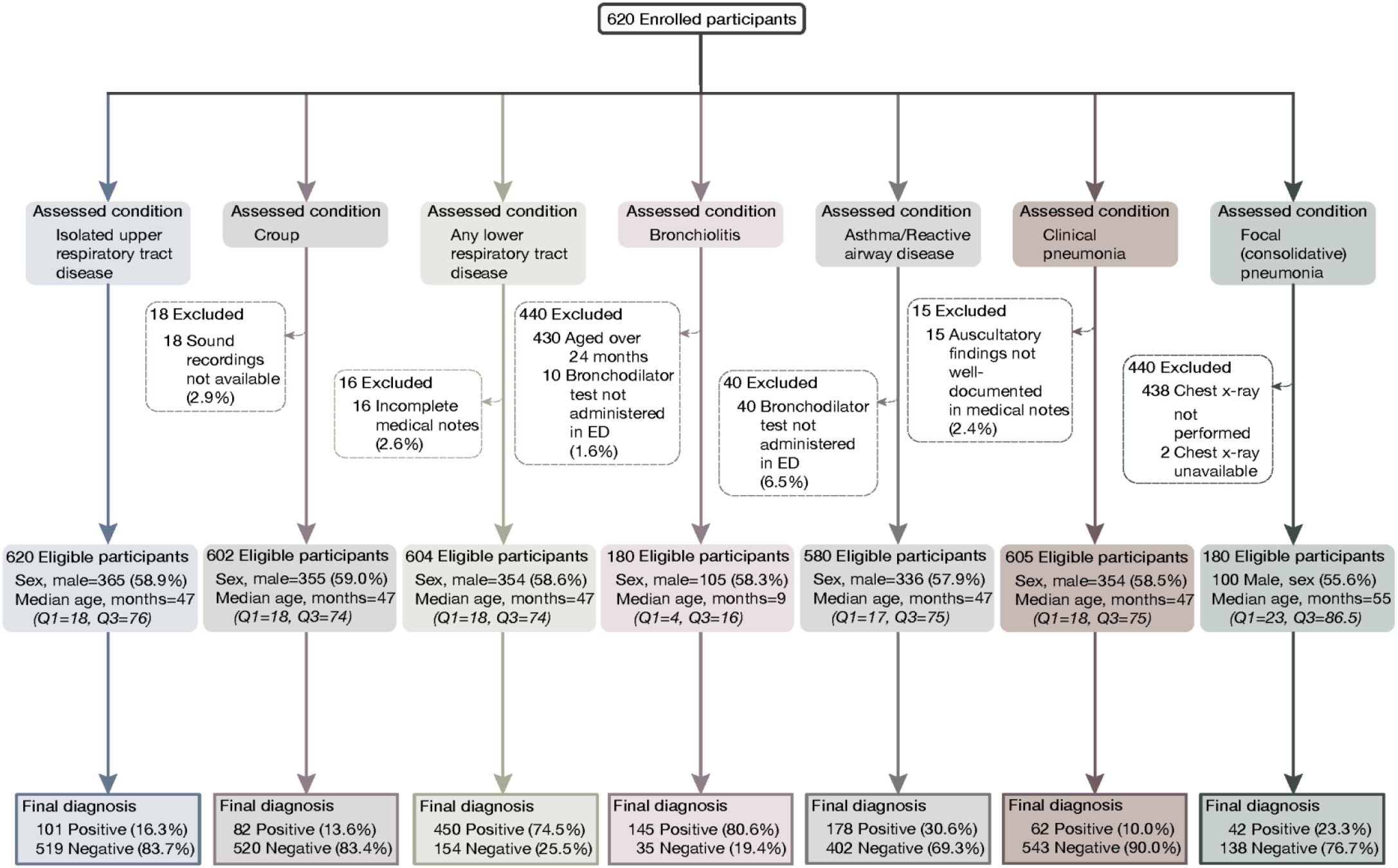
Participant demographics and flow through the study by individual study diseases.

### Comparison between the emergency department and expert panel diagnoses

The PPA and NPA between E.D. and panel diagnoses are shown in Table 2, along with the number of missed cases (false negative) and false-alarm cases (false positive). There were no differences between the two study sites.

**Table 2.** Agreement of E.D. diagnosis with the expert panel consensus diagnosis.

| Final Diagnosis | PPA<sup>a</sup> with expert panel<sup>b</sup> diagnosis (% [95% CI]) | NPA<sup>c</sup> with expert panel diagnosis (% [95% CI]) | Missed cases (n; FNR<sup>d</sup>) | ED diagnosis of false-negative cases (n) | False alarms (n; FPR<sup>e</sup>) | Expert panel diagnosis of false-positive cases<sup>f</sup> |
| --- | --- | --- | --- | --- | --- | --- |
| Isolated URTD | 61.4<br>[51.2, 70.9] | 90.9<br>[88.1, 93.3] | 39/101<br>(38.6) | - No respiratory disease (12)<br>- Croup (14)<br>- LRTD (15)<br>- Bronchiolitis (1)<br>- Asthma/RAD (9)<br>- Clinical pneumonia (1) | 47/51<br>9 (9.1) | - Croup (11)<br>- LRTD (37)<br>- Bronchiolitis (13)<br>- Asthma/RAD (3)<br>- Clinical pneumonia (6)<br>- Focal (consolidative) pneumonia (3) |
| Croup | 75.6<br>[64.9, 84.4] | 97.9<br>[96.2, 98.9] | 20/82<br>(24.4) | - iURTD (11)<br>- LRTD (8)<br>- Bronchiolitis (2)<br>- Asthma/RAD (4) | 11/52<br>0 (2.1) | - iURTD (6)<br>- LRTD (5)<br>- Bronchiolitis (3)<br>- Clinical pneumonia (1)<br>- Focal (consolidative) pneumonia (1) |
| LRTD | 86.4<br>[82.9, 89.5] | 92.9<br>[87.6, 96.4] | 61/450<br>(13.6) | - No respiratory disease (18)<br>- iURTD (37)<br>- Croup (6) | 11/15<br>4 (7.1) | - iURTD (9)<br>- Croup (2) |
| Bronchiolitis | 66.9<br>[58.6, 74.5] | 94.3<br>[80.8, 99.3] | 48 /145<br>(33.1) | - No respiratory disease (3)<br>- iURTD (14)<br>- Croup (3)<br>- LRTD (not bronchiolitis) (29)<br>- Asthma/RAD (13)<br>- Clinical pneumonia (12)<br>- Focal (consolidative) pneumonia (7) | 2/35<br>(5.7) | - Croup (1)<br>- LRTD (1)<br>- Clinical pneumonia (1)<br>- Focal (consolidative) pneumonia (1) |
| Asthma/RAD | 91.0<br>[85.8, 94.8] | 93.0<br>[90.1, 95.3] | 16/178<br>(9.0) | - No respiratory disease (1)<br>- iURTD (2)<br>- Croup (1)<br>- LRTD (not asthma) (12)<br>- Bronchiolitis (4)<br>- Clinical pneumonia (7)<br>- Focal (consolidative) pneumonia (2) | 28/40<br>2 (7.0) | - iURTD (6)<br>- LRTD (22)<br>- Bronchiolitis (12)<br>- Clinical pneumonia (3)<br>- Focal (consolidative) pneumonia (3) |
| Clinical pneumonia | 62.9<br>[49.7, 74.8] | 95.0<br>[92.8, 96.7] | 23/62<br>(37.1) | - No respiratory disease (9)<br>- iURTD (6)<br>- Croup (1)<br>- LRTD (not pneumonia) (8)<br>- Bronchiolitis (2)<br>- Asthma/RAD (2)<br>- Focal (consolidative) pneumonia (1) | 27/54<br>3 (5.0) | - iURTD (1)<br>- LRTD (26)<br>- Bronchiolitis (11)<br>- Asthma/RAD (7) |
| Focal (consolidative) pneumonia | 54.8<br>[38.7, 70.2] | 86.2<br>[79.3, 91.5] | 19/42<br>(45.2) | - No respiratory disease (6)<br>- iURTD (3)<br>- Croup (1)<br>- LRTD (not pneumonia) (10)<br>- Bronchiolitis (3)<br>- Asthma/RAD (1)<br>- Clinical pneumonia (5) | 19/13<br>8<br>(13.7) | - LRTD (19)<br>- Asthma/RAD (2)<br>- Clinical pneumonia (7) |

<sup>a</sup>PPA: Positive Percent Agreement
<sup>b</sup>EP: Expert panel
<sup>c</sup>NPA: negative percent agreement
<sup>d</sup>FNR: False-negative rate
<sup>e</sup>FPR: False-positive rate
PPA and NPA data are presented as percentage and 95% confidence intervals. False-positive and false-negative data presented as absolute ratio and percentages of condition negative and positive, respectively. Numbers do not equal total missed or total false alarm cases as more than one study diagnosis could be allocated for each participant

Asthma/RAD was the most reliably diagnosed condition (91% correctly identified as positive) followed by LRTD (86.4%), croup (75.6%), bronchiolitis (66.9%) and iURTD (61.4%). Pneumonia was the least reliably diagnosed condition, with clinical pneumonia identified in 62.9% of participants, while focal (consolidative) pneumonia was correctly identified in only 54.8% of participants. The NPA for all study diseases was higher than the PPA.

We explored the missed and false alarm cases by study disease, including the diagnoses erroneously attributed to each participant (Table 2). There were common errors with differentiating asthma/RAD from bronchiolitis, croup from iURTD, and iURTD from LRTD. In 10 out of 42 cases of focal (consolidative) pneumonia, the E.D. did not detect any type of LRTD, and in six instances, no respiratory disease was detected. E.D. clinicians diagnosed the presence of a LRTD (10/19) or a wheezy condition (4/19) when they missed focal (consolidative) pneumonia.

Chest imaging was performed on 153/620 (24.6%) participants in the E.D. The E.D. missed 16/39 (41%) cases of radiologically confirmed consolidation and over-diagnosed the presence of consolidation in 19/138 (13.7%) of non-pneumonia cases. Many children without pneumonia had chest x-rays, not in accordance with published guidelines: iURTI (9/101, 8.9%), croup (3/82, 3.6%), bronchiolitis (22/143, 15.4%) and asthma/RAD (31/177, 17.5%).

Children frequently had inappropriate tests and treatment in the ED. Participants with isolated URTD (n=101) had bronchodilator tests (n=8, 7.9%), blood cultures (n=2, 2%), viral nasal swabs (n=6, 5.9%) and antibiotics (n=7, 6.9%). Participants with bronchiolitis (n=145) had blood cultures (n=20, 13.8%) and viral nasal swabs (n=95, 65.5%) and antibiotics (n=21, 14.5%). In children with asthma, 13/178 (7.3%) were prescribed antibiotics unnecessarily.

## DISCUSSION

Diagnostic errors cost health systems billions of dollars each year and result in a notable amount of preventable harm. Our findings show that acute respiratory disorders are frequently misdiagnosed in optimally resourced E.D.s despite them being the most common reason for presentation. While asthma (91%) and LRTDs (86%) were well identified, over 45% of focal (consolidative) pneumonia, 39% of iURTD, 25% of croup and 33% of bronchiolitis cases were missed. Each diagnostic error was compounded by participants receiving an incorrect diagnosis, including 8% of LRTDs being assigned as iURTD and 14% of focal (consolidative) pneumonias classified as “no respiratory disease”.

In the assessment of respiratory disorders, the first decision point is the distinction of isolated upper airway from lower respiratory tract diseases such as pneumonia and asthma. Doctors are more likely to prioritize assessment and treatment for LRTDs as these conditions tend to be more severe. In our study, E.D. clinicians correctly identified a LRTD in 86% of cases, however then had trouble differentiating the more specific diagnoses of bronchiolitis and pneumonia. In addition,15% of children with iURTDs were misdiagnosed as having a LRTD. Although this overly cautious approach could be considered safe, it does result in unnecessary investigations and treatments which carry adverse consequences. Many study participants who did not have pneumonia received unnecessary antibiotics (6.9%–14.5%) and x-rays (3.6%–17.5%), resulting in poor antibiotic stewardship and ionizing radiation exposure. Economically, the annual cost of such defensive medicine and over-investigation to guard against malpractice lawsuits is estimated to be at least USD 25–60 billion (30).

Due to overlapping features, it can be challenging for clinicians to differentiate between conditions that present with acute wheeze such as bronchiolitis and asthma. This is even more complicated when assessing young children who cannot always communicate their symptoms. In order to differentiate between wheezy conditions, physicians rely on bronchodilator tests and professional experience. In resource-poor areas where bronchodilator testing is not usually done, up to 50% of children under the age of five years who are diagnosed with pneumonia using WHO/Integrated management of childhood illness guidelines could subsequently be reclassified as having asthma (31-33). In our study, the accurate identification of asthma was excellent (91% of cases) except in younger children (<24 months) in which 9% of bronchiolitis cases were misdiagnosed as asthma. These diagnostic errors may have resulted from individual clinicians’ interpretation of bronchodilator tests.

The accurate diagnosis of pneumonia is dependent on the availability of clinical expertise and diagnostic support resources (34-37). Without radiology, clinicians must rely on auscultatory and clinical findings. However, the diagnosis of community-acquired pneumonia using presenting clinical features alone is often inadequate, with primary care providers missing up to 71% of cases in adults (38-40). In our definition of clinical pneumonia, we included patients with generalized auscultatory changes along with patients with only focal changes to capture viral, atypical (mycoplasma) and bacterial infections. This approach can increase the possibility of misdiagnosis, as these features are also associated with conditions such as asthma and bronchiolitis. Viral and atypical chest infections typically have generalized signs, while bacterial infections are typically more localized, but there is considerable overlap between groups. Using this broad definition of pneumonia, we found that E.D. clinicians missed 37% of cases. Nine of the missed cases (n=23) were instead diagnosed with “no respiratory disease” while six were diagnosed with an isolated URTD. Conversely, 96% of patients erroneously diagnosed with clinical pneumonia had another LRTD, including 26% who had asthma and did not receive appropriate treatment.

The most frequently missed condition in the study was focal (consolidative) pneumonia (PPA 55%, NPA 86%), which was also the most potentially serious condition because of probable bacterial etiology. As early detection and commencement of antibiotic therapy is essential for best outcomes these errors represent a serious cause of preventable harm. Globally, pneumonia is the leading cause of childhood death under five years of age, with the highest mortality rates seen in low-resourced areas (41). Although our study sites had access to specialist trained pediatric doctors and full diagnostic imaging support, they missed nearly one in two cases of focal (consolidative) pneumonia. In comparison, the symptom-based algorithm developed by the WHO/IMCD to detect pneumonia in low-resourced settings without radiology or expert healthcare has been found to have a similar sensitivity of 67% but a lower specificity of 60% to our study when compared to diagnosis by lung ultrasound (42). A 2021 study in Tanzania found that the algorithm had a much lower diagnostic sensitivity of 25%, and in a Canadian study using a population more comparable to our study, the positive and negative predictive values were also 25% (43, 44). Studies in adult patients with suspected community-acquired pneumonia have reported higher false-positive rates than our results. A study across three hospitals that included 800 patients admitted from the E.D. with a community-acquired pneumonia diagnosis revealed 219 (27%) had a non-pneumonia diagnosis upon discharge (45). In another, a cohort of 195 adult patients admitted to a hospital ward with an E.D. diagnosis of pneumonia included 18% with normal chest x-rays (46). In addition, 29% of patients had a discordant discharge diagnosis, with 54% of these patients being diagnosed with an upper respiratory infection and 30% classified as having no infection at all.

We found a substantial error rate in the interpretation of chest x-rays. Only 59% of x-rays with consolidation were correctly diagnosed in E.D., while 14% of normal x-rays were erroneously thought to have consolidation. This error was a significant contributing factor to the poor diagnostic performance for focal (consolidative) pneumonia in our study. Difficulties associated with identifying pneumonia and consolidation on x-ray have been described in the adult literature, including poor interrater agreement (34-37) between E.D. doctors and radiologists. Lung ultrasound examination has emerged as an effective tool. When used by trained operators a meta-analysis found a pooled sensitivity of 95% and a specificity of 95%. Diagnostic error rates for pneumonia could be reduced by having radiologists work with E.D. teams in real-time, but this approach would be resource-intensive and not possible in small or regional hospitals.

Diagnostic errors are often caused by human factors such as cognition, tiredness, distraction and miscommunication (47). Emergency departments are by nature distracting environments where clinicians are required to see multiple patients concurrently, often while tired. Strategies to lessen these influences have led to the development of objective digital technologies using artificial intelligence (A.I.) and machine learning. AI-based systems which mimic the clinical diagnostic process have shown some promising results. A system used to interrogate electronic medical records has been reported to accurately identify many pediatric conditions and differentiate upper from lower respiratory disease with over 87% accuracy (48). This approach, however, relies on the quality and generalizability of the training data sets and can only be used after all information has been entered into the patient’s medical record. Studies of algorithms that use point-of-care automated cough-centered analysis have reported good diagnostic accuracy for respiratory diseases, including pneumonia, correctly identifying 87% of children and 86% of adults without the need for clinical examination or investigations (49-52). In settings with limited resources, such algorithms might provide a diagnostic method equal to the accuracy of well-resourced E.D.s. The development of automated algorithms to detect pneumonia based on ultrasound patterns has also shown promise, potentially reducing the need for trained operators and chest x-rays (53).

## Limitations

We were confronted with several challenges during the design of our study. As some of the study diseases do not have clear objective diagnostic markers or tests, we articulated strict definitions for the adjudication panel to use based on internationally published criteria (21-25). We have found no other studies that have used reference diagnostic criteria as stringent as this. There were limitations relating to wheezy conditions, such as asthma, in which an accurate diagnosis relies on bronchodilator test responsiveness. However, unless formal lung function tests are conducted, interpretation of the response to bronchodilators is subjective (54). As it is not feasible to perform lung function tests in young children in E.D., nor is it recommended for children younger than six, we acknowledge that the interpretation of bronchodilator tests in our study was subjective. However, this is the current standard of care in clinical practice. To improve accuracy, we insisted a bronchodilator test be administered in the ED and the response recorded before assessing for either asthma or bronchiolitis. For children aged 1-2 years, the differentiation between bronchiolitis and asthma depends on bronchodilator test response, with bronchiolitis patients being unresponsive. We excluded cases (n=10) from our analysis if bronchodilator tests were not done to avoid diagnostic classification errors.

To minimize the over-diagnosis of focal (consolidative) pneumonia we defined a study group to reflect focal, lobar pathology by requiring a specialist radiologist to report chest x-rays and ensured the absence of any generalized wheezy conditions. For this group, we used the radiological definitions developed by the WHO to detect bacterial lung disease in post-vaccination surveillance programs (28). We adopted this approach to avoid diagnosing focal (consolidative) pneumonia in cases of generalized LRTDs, such as asthma, bronchiolitis, and pneumonitis, when radiology was inappropriately performed. Assessment guidelines recommend against diagnostic imaging in bronchiolitis and asthma (21,26,55). Despite this, chest x-rays were inappropriately ordered in our study for these conditions, resulting in unnecessary radiation and antibiotic use when pneumonia was mistakenly diagnosed (56-59).

We used a panel of three experts to provide a diagnostic consensus. Although this is a non-reference standard and one that is not attainable in real-life clinical practice, it provided us with the best achievable diagnostic standard. Studies that have used a single reviewer have been found to lack validity, and even those using two reviewers have shown low agreement (60-63). Other studies have reported little benefit in using more than three assessors, and many studies have only engaged a single or second independent reviewer (11,64).

An analysis of diagnostic error studies that rely on chart review shows that the clinical data needed to make diagnoses are often missing in cases where an error has been made (65,66). Our panel, who did not have an opportunity to examine the patients or order tests, relied on medical charts and cough recordings. As the E.D. and panel recorded their diagnoses using the same examination findings, this could have led to underestimating the true frequency of diagnostic errors.

## Conclusion

To the best of our knowledge, this is the first study to report on diagnostic error rates in well-resourced E.D.s for undifferentiated acute childhood respiratory diseases. Although these conditions represent the most common reasons for children to be taken to an ED and account for some of the more serious pediatric disorders, they are frequently misdiagnosed. The high diagnostic error rate for pneumonia is particularly concerning. Further, our results point to the risks posed to individuals caused by these errors, including the prescription of inappropriate tests and treatments whilst appropriate therapy is delayed or not given. More specifically, study participants received unnecessary antibiotics for bronchiolitis (14.5%), asthma (7.3%) and iURTD (6.9%) and were subjected to ionizing radiation. Incorrect antibiotic use carries implications for both individuals and the broader community in terms of antibiotic resistance and resource allocation.

To reduce the risk of harm associated with diagnostic errors, further studies should focus on objective methods to improve diagnostic accuracy, such as AI-based systems. Repeating this study in environments with limited clinical and diagnostic support resources would help to establish a baseline from which to assess the usefulness of new diagnostic modalities.

## Data Availability

The datasets supporting the conclusion of this article are available on reasonable request from PP after reciprocal ethical approval has been obtained. The cough recordings are not available but will be uploaded as an educational tool at the conclusion of the Breathe Easy development program in 2022.

## References

1. Committee on Diagnostic Error in Health Care, Board on Health Care Services, Institute of Medicine, et al. In: Balogh EP, Miller BT, Ball JR, eds. Improving Diagnosis in Health Care. Washington (DC): National Academies Press (US) 2015.

2. Medford-Davis LN, Singh H, Mahajan P. Diagnostic Decision-Making in the Emergency Department. Pediatr Clin North Am 2018;65(6):1097–105. doi: 10.1016/j.pcl.2018.07.003 [published Online First: 2018/11/18]

3. Zwaan L, Singh H. The challenges in defining and measuring diagnostic error. Diagnosis (Berl) 2015;2(2):97–103. doi: 10.1515/dx-2014-0069 [published Online First: 2016/03/10]

4. Newman-Toker DE, Pronovost PJ. Diagnostic errors--the next frontier for patient safety. JAMA 2009;301(10):1060–2. doi: 10.1001/jama.2009.249 [published Online First: 2009/03/13]

5. Meyer AN, Payne VL, Meeks DW, et al. Physicians’ diagnostic accuracy, confidence, and resource requests: a vignette study. JAMA Intern Med 2013;173(21):1952–8. doi: 10.1001/jamainternmed.2013.10081 [published Online First: 2013/08/28]

6. United Nations Children’s Fund. Levels and trends in child mortality, estimates developed by the UN inter-agency group for child mortality estimation. NewYork, 2014.

7. Warrick C, Patel P, Hyer W, et al. Diagnostic error in children presenting with acute medical illness to a community hospital. Int J Qual Health Care 2014;26(5):538–46. doi: 10.1093/intqhc/mzu066 [published Online First: 2014/07/09]

8. Perrem LM, Fanshawe TR, Sharif F, et al. A national physician survey of diagnostic error in paediatrics. Eur J Pediatr 2016;175(10):1387–92. doi: 10.1007/s00431-016-2772-0 [published Online First: 2016/09/16]

9. Singh H, Thomas EJ, Wilson L, et al. Errors of diagnosis in pediatric practice: a multisite survey. Pediatrics 2010;126(1):70–9. doi: 10.1542/peds.2009-3218 [published Online First: 2010/06/23]

10. Newman-Toker DE, Makary MA. Measuring diagnostic errors in primary care: the first step on a path forward. Comment on “Types and origins of diagnostic errors in primary care settings”. JAMA Intern Med 2013;173(6):425–6. doi: 10.1001/jamainternmed.2013.225 [published Online First: 2013/02/27]

11. Singh H, Giardina TD, Meyer AN, et al. Types and origins of diagnostic errors in primary care settings. JAMA Intern Med 2013;173(6):418–25. doi: 10.1001/jamainternmed.2013.2777 [published Online First: 2013/02/27]

12. Rubin G, Meyer AND. Diagnostic errors and harms in primary care: insights to action. BMJ Qual Saf 2021 doi: 10.1136/bmjqs-2020-012423 [published Online First: 2021/06/02]

13. Andel C, Davidow SL, Hollander M, et al. The economics of health care quality and medical errors. J Health Care Finance 2012;39(1):39–50. [published Online First: 2012/11/20]

14. Selbst SM, Friedman MJ, Singh SB. Epidemiology and etiology of malpractice lawsuits involving children in US emergency departments and urgent care centers. Pediatr Emerg Care 2005;21(3):165–9. [published Online First: 2005/03/04]

15. McDermott KW, Stocks C, Wj F. Overview of Pediatric Emergency Department Visits, 2015. HCUP Statistical Brief #242. Rockville, MD: Agency for Healthcare Research and Quality, August 2018.

16. Ostergaard MS, Nantanda R, Tumwine JK, et al. Childhood asthma in low income countries: an invisible killer? Prim Care Respir J 2012;21(2):214–9. doi: 10.4104/pcrj.2012.00038 [published Online First: 2012/05/25]

17. Elphick HE, Lancaster GA, Solis A, et al. Validity and reliability of acoustic analysis of respiratory sounds in infants. Arch Dis Child 2004;89(11):1059–63. doi: 10.1136/adc.2003.046458 [published Online First: 2004/10/23]

18. McCollum ED, Park DE, Watson NL, et al. Listening panel agreement and characteristics of lung sounds digitally recorded from children aged 1-59 months enrolled in the Pneumonia Etiology Research for Child Health (PERCH) case-control study. BMJ Open Respir Res 2017;4(1):e000193. doi: 10.1136/bmjresp-2017-000193 [published Online First: 2017/09/09]

19. Looijmans-van den Akker I, van Luijn K, Verheij T. Overdiagnosis of asthma in children in primary care: a retrospective analysis. Br J Gen Pract 2016;66(644):e152–7. doi: 10.3399/bjgp16X683965 [published Online First: 2016/02/27]

20. Yang CL, Simons E, Foty RG, et al. Misdiagnosis of asthma in schoolchildren. Pediatr Pulmonol 2017;52(3):293–302. doi: 10.1002/ppul.23541 [published Online First: 2016/08/10]

21. O’Brien S, Borland ML, Cotterell E, et al. Australasian bronchiolitis guideline. J Paediatr Child Health 2019;55(1):42–53. doi: 10.1111/jpc.14104 [published Online First: 2018/07/17]

22. Perth Children’s Hospital. Emergency Department Guidelines - Pneumonia 2018 [Available from: https://pch.health.wa.gov.au/For-health-professionals/Emergency-Department-Guidelines/Pneumonia.

23. Perth Children’s Hospital. Emergency Department Guidelines - Cough 2018 [Available from: https://pch.health.wa.gov.au/For-health-professionals/Emergency-Department-Guidelines/Cough.

24. Perth Children’s Hospital. Emergency Department Guidelines - Asthma 2018 [Available from: https://pch.health.wa.gov.au/For-health-professionals/Emergency-Department-Guidelines/Asthma.

25. National Institute for Health and Care Excellence (NICE). Asthma: diagnosis, monitoring and chronic asthma management. NICE Guideline, 2017.

26. National Institute for Health and Care Excellence (NICE). Bronchiolitis in children: diagnosis and management. NICE Guideline, 2015.

27. Bradley JS, Byington CL, Shah SS, et al. The management of community-acquired pneumonia in infants and children older than 3 months of age: clinical practice guidelines by the Pediatric Infectious Diseases Society and the Infectious Diseases Society of America. Clin Infect Dis 2011;53(7):e25–76. doi: 10.1093/cid/cir531 [published Online First: 2011/09/02]

28. Cherian T, Mulholland EK, Carlin JB, et al. Standardized interpretation of paediatric chest radiographs for the diagnosis of pneumonia in epidemiological studies. Bull World Health Organ 2005;83(5):353–9. doi: S0042-96862005000500011 [published Online First: 2005/06/25]

29. World Health Organization Pneumonia Vaccine Trial Investigators’ Group. Standardization of interpretation of chest radiographs for the diagnosis of pneumonia in children. Geneva, 2001.

30. Mello MM, Chandra A, Gawande AA, et al. National costs of the medical liability system. Health Aff (Millwood) 2010;29(9):1569–77. doi: 10.1377/hlthaff.2009.0807 [published Online First: 2010/09/08]

31. Nantanda R, Tumwine JK, Ndeezi G, et al. Asthma and pneumonia among children less than five years with acute respiratory symptoms in Mulago Hospital, Uganda: evidence of under-diagnosis of asthma. PLoS One 2013;8(11):e81562. doi: 10.1371/journal.pone.0081562 [published Online First: 2013/12/07]

32. Sachdev HP, Mahajan SC, Garg A. Improving antibiotic and bronchodilator prescription in children presenting with difficult breathing: experience from an urban hospital in India. Indian Pediatr 2001;38(8):827–38. [published Online First: 2001/08/25]

33. Hazir T, Qazi S, Nisar YB, et al. Assessment and management of children aged 1-59 months presenting with wheeze, fast breathing, and/or lower chest indrawing; results of a multicentre descriptive study in Pakistan. Arch Dis Child 2004;89(11):1049–54. doi: 10.1136/adc.2003.035741 [published Online First: 2004/10/23]

34. Bada C, Carreazo NY, Chalco JP, et al. Inter-observer agreement in interpreting chest X-rays on children with acute lower respiratory tract infections and concurrent wheezing. Sao Paulo Med J 2007;125(3):150–4. doi: 10.1590/s1516-31802007000300005 [published Online First: 2007/10/10]

35. Niederman MS. Review of treatment guidelines for community-acquired pneumonia. Am J Med 2004;117 Suppl 3A:51S–57S. doi: 10.1016/j.amjmed.2004.07.008 [published Online First: 2004/09/14]

36. Washington L, Palacio D. Imaging of bacterial pulmonary infection in the immunocompetent patient. Semin Roentgenol 2007;42(2):122–45. doi: 10.1053/j.ro.2006.08.008 [published Online First: 2007/03/31]

37. Waterer GW. The Diagnosis of Community-acquired Pneumonia. Do We Need to Take a Big Step Backward? Am J Respir Crit Care Med 2015;192(8):912–3. doi: 10.1164/rccm.201507-1460ED [published Online First: 2015/10/16]

38. van Vugt SF, Verheij TJ, de Jong PA, et al. Diagnosing pneumonia in patients with acute cough: clinical judgment compared to chest radiography. Eur Respir J 2013;42(4):1076–82. doi: 10.1183/09031936.00111012 [published Online First: 2013/01/26]

39. Arts L, Lim EHT, van de Ven PM, et al. The diagnostic accuracy of lung auscultation in adult patients with acute pulmonary pathologies: a meta-analysis. Sci Rep 2020;10(1):7347. doi: 10.1038/s41598-020-64405-6 [published Online First: 2020/05/02]

40. Htun TP, Sun Y, Chua HL, et al. Clinical features for diagnosis of pneumonia among adults in primary care setting: A systematic and meta-review. Sci Rep 2019;9(1):7600. doi: 10.1038/s41598-019-44145-y [published Online First: 2019/05/22]

41. Liu L, Oza S, Hogan D, et al. Global, regional, and national causes of child mortality in 2000-13, with projections to inform post-2015 priorities: an updated systematic analysis. Lancet 2015;385(9966):430–40. doi: 10.1016/S0140-6736(14)61698-6 [published Online First: 2014/10/05]

42. Chavez MA, Naithani N, Gilman RH, et al. Agreement Between the World Health Organization Algorithm and Lung Consolidation Identified Using Point-of-Care Ultrasound for the Diagnosis of Childhood Pneumonia by General Practitioners. Lung 2015;193(4):531–8. doi: 10.1007/s00408-015-9730-x [published Online First: 2015/04/30]

43. Salisbury T, Redfern A, Fletcher EK, et al. Correct diagnosis of childhood pneumonia in public facilities in Tanzania: a randomised comparison of diagnostic methods. BMJ Open 2021;11(5):e042895. doi: 10.1136/bmjopen-2020-042895 [published Online First: 2021/05/26]

44. Rothrock SG, Green SM, Fanelli JM, et al. Do published guidelines predict pneumonia in children presenting to an urban ED? Pediatr Emerg Care 2001;17(4):240–3. doi: 10.1097/00006565-200108000-00003 [published Online First: 2001/08/09]

45. Chandra A, Nicks B, Maniago E, et al. A multicenter analysis of the ED diagnosis of pneumonia. Am J Emerg Med 2010;28(8):862–5. doi: 10.1016/j.ajem.2009.04.014 [published Online First: 2010/10/05]

46. Atamna A, Shiber S, Yassin M, et al. The accuracy of a diagnosis of pneumonia in the emergency department. Int J Infect Dis 2019;89:62–65. doi: 10.1016/j.ijid.2019.08.027 [published Online First: 2019/09/04]

47. World Health Organization. Diagnostic Errors: Technical Series on Safer Primary Care. Geneva, 2016.

48. Liang H, Tsui BY, Ni H, et al. Evaluation and accurate diagnoses of pediatric diseases using artificial intelligence. Nat Med 2019;25(3):433–38. doi: 10.1038/s41591-018-0335-9 [published Online First: 2019/02/12]

49. Porter P, Abeyratne U, Swarnkar V, et al. A prospective multicentre study testing the diagnostic accuracy of an automated cough sound centred analytic system for the identification of common respiratory disorders in children. Respir Res 2019;20(1):81. doi: 10.1186/s12931-019-1046-6 [published Online First: 2019/06/07]

50. Porter P, Brisbane J, Abeyratne U, et al. Diagnosing community-acquired pneumonia via a smartphone-based algorithm: a prospective cohort study in primary and acute-care consultations. Br J Gen Pract 2021;71(705):e258–e65. doi: 10.3399/BJGP.2020.0750 [published Online First: 2021/02/10]

51. Porter P, Claxton S, Brisbane J, et al. Diagnosing Chronic Obstructive Airway Disease on a Smartphone Using Patient-Reported Symptoms and Cough Analysis: Diagnostic Accuracy Study. JMIR Form Res 2020;4(11):e24587. doi: 10.2196/24587 [published Online First: 2020/11/11]

52. Abeyratne UR, Swarnkar V, Setyati A, et al. Cough sound analysis can rapidly diagnose childhood pneumonia. Ann Biomed Eng 2013;41(11):2448–62. doi: 10.1007/s10439-013-0836-0 [published Online First: 2013/06/08]

53. Correa M, Zimic M, Barrientos F, et al. Automatic classification of pediatric pneumonia based on lung ultrasound pattern recognition. PLoS One 2018;13(12):e0206410. doi: 10.1371/journal.pone.0206410 [published Online First: 2018/12/06]

54. Nielsen KG, Bisgaard H. Discriminative capacity of bronchodilator response measured with three different lung function techniques in asthmatic and healthy children aged 2 to 5 years. Am J Respir Crit Care Med 2001;164(4):554–9. doi: 10.1164/ajrccm.164.4.2006119 [published Online First: 2001/08/25]

55. Ralston SL, Lieberthal AS, Meissner HC. Ralston SL, Lieberthal AS, Meissner HC, et al. Clinical Practice Guideline: The Diagnosis, Management, and Prevention of Bronchiolitis. Pediatrics. 2014;134(5):e1474-e1502. Pediatrics 2015;136(4):782. doi: 10.1542/peds.2015-2862 [published Online First: 2015/10/03]

56. Florin TA, Byczkowski T, Ruddy RM, et al. Variation in the management of infants hospitalized for bronchiolitis persists after the 2006 American Academy of Pediatrics bronchiolitis guidelines. J Pediatr 2014;165(4):786–92 e1. doi: 10.1016/j.jpeds.2014.05.057 [published Online First: 2014/07/13]

57. Johnson LW, Robles J, Hudgins A, et al. Management of bronchiolitis in the emergency department: impact of evidence-based guidelines? Pediatrics 2013;131 Suppl 1:S103–9. doi: 10.1542/peds.2012-1427m [published Online First: 2013/03/15]

58. Narayanan S, Magruder T, Walley SC, et al. Relevance of chest radiography in pediatric inpatients with asthma. J Asthma 2014;51(7):751–5. doi: 10.3109/02770903.2014.909459 [published Online First: 2014/03/29]

59. American Academy of Pediatrics Subcommittee on D, Management of B. Diagnosis and management of bronchiolitis. Pediatrics 2006;118(4):1774–93. doi: 10.1542/peds.2006-2223 [published Online First: 2006/10/04]

60. Gunderson CG, Bilan VP, Holleck JL, et al. Prevalence of harmful diagnostic errors in hospitalised adults: a systematic review and meta-analysis. BMJ Qual Saf 2020;29(12):1008–18. doi: 10.1136/bmjqs-2019-010822 [published Online First: 2020/04/10]

61. Forster AJ, O’Rourke K, Shojania KG, et al. Combining ratings from multiple physician reviewers helped to overcome the uncertainty associated with adverse event classification. J Clin Epidemiol 2007;60(9):892–901. doi: 10.1016/j.jclinepi.2006.11.019 [published Online First: 2007/08/11]

62. Zwaan L, de Bruijne M, Wagner C, et al. Patient record review of the incidence, consequences, and causes of diagnostic adverse events. Arch Intern Med 2010;170(12):1015–21. doi: 10.1001/archinternmed.2010.146 [published Online First: 2010/06/30]

63. Singh H, Giardina TD, Forjuoh SN, et al. Electronic health record-based surveillance of diagnostic errors in primary care. BMJ Qual Saf 2012;21(2):93–100. doi: 10.1136/bmjqs-2011-000304 [published Online First: 2011/10/15]

64. Kahan BC, Feagan B, Jairath V. A comparison of approaches for adjudicating outcomes in clinical trials. Trials 2017;18(1):266. doi: 10.1186/s13063-017-1995-3 [published Online First: 2017/06/10]

65. Newman-Toker DE. Charted records of dizzy patients suggest emergency physicians emphasize symptom quality in diagnostic assessment. Ann Emerg Med 2007;50(2):204–5. doi: 10.1016/j.annemergmed.2007.03.037 [published Online First: 2007/07/24]

66. Kerber KA, Morgenstern LB, Meurer WJ, et al. Nystagmus assessments documented by emergency physicians in acute dizziness presentations: a target for decision support? Acad Emerg Med 2011;18(6):619–26. doi: 10.1111/j.1553-2712.2011.01093.x [published Online First: 2011/06/17]

